# Comparison of Mental Health Symptom Changes from pre-COVID-19 to COVID-19 by Sex or Gender: A Systematic Review and Meta-analysis

**DOI:** 10.1101/2021.06.28.21259384

**Authors:** Tiffany Dal Santo, Ying Sun, Yin Wu, Chen He, Yutong Wang, Xiaowen Jiang, Kexin Li, Olivia Bonardi, Ankur Krishnan, Jill T. Boruff, Danielle B. Rice, Sarah Markham, Brooke Levis, Marleine Azar, Dipika Neupane, Amina Tasleem, Anneke Yao, Ian Thombs-Vite, Branka Agic, Christine Fahim, Michael S. Martin, Sanjeev Sockalingam, Gustavo Turecki, Andrea Benedetti, Brett D. Thombs

## Abstract

**Importance:** Women and gender-diverse individuals have faced disproportionate socioeconomic burden during COVID-19. There have been reports that this has translated into greater negative changes in mental health, but this has been based on cross-sectional research that has not accounted for pre-COVID-19 differences.

**Objective:** To compare mental health symptom changes since pre-COVID-19 by sex or gender.

**Data Sources:** MEDLINE, PsycINFO, CINAHL, EMBASE, Web of Science, China National Knowledge Infrastructure, Wanfang, medRxiv, and Open Science Framework (December 31, 2019 to August 30, 2021).

**Study Selection:** Eligible studies compared mental health symptom changes from pre-COVID-19 to COVID-19 by sex or gender.

**Data Extraction and Synthesis:** Data was extracted by a single reviewer with validation by a second reviewer. Adequacy of study methods and reporting was assessed using an adapted version of the Joanna Briggs Institute Checklist for Prevalence Studies. A restricted maximum-likelihood random-effects meta-analyses was conducted.

**Main Outcomes and Measures:** Anxiety symptoms, depression symptoms, general mental health, and stress measured continuously or dichotomously.

**Results:** 12 studies (10 unique cohorts) were included. All compared females or women to males or men; none included gender-diverse individuals. Continuous symptom change differences were not statistically significant for depression (standardized mean difference [SMD]= 0.12, 95% CI -0.09 to 0.33; 4 studies, 4,475 participants; I^2^=69.0%) and stress (SMD= - 0.10, 95% CI -0.21 to 0.01; 4 studies, 1,533 participants; I^2^=0.0%), but anxiety (SMD= 0.15, 95% CI 0.07 to 0.22; 4 studies, 4,344 participants; I^2^=3.0%) and general mental health (SMD= 0.15, 95% CI 0.12 to 0.18; 3 studies, 15,692 participants; I^2^=0.0%) worsened more among females or women than males or men during COVID-19. There were no significant differences in changes in proportion above a cut-off: anxiety (difference= -0.05, 95% CI -0.20 to 0.11; 1 study, 217 participants), depression (difference= 0.12, 95% CI -0.03 to 0.28; 1 study, 217 participants), general mental health (difference= -0.03, 95% CI -0.09 to 0.04; 3 studies, 18,985 participants; I^2^=94.0%), stress (difference= 0.04, 95% CI -0.10 to 0.17; 1 study, 217 participants).

**Conclusion and Relevance:** Mental health outcomes did not differ or were worse by amounts below thresholds for clinical significance for women compared to men.

**Registration:** PROSPERO (CRD42020179703).

**KEY MESSAGES:** *Question:* Did mental health symptoms worsen more for females or women than males or men in COVID-19?

*Findings:* We reviewed almost 65,000 citations and identified 12 studies that provided data to directly compare mental health symptom changes from pre-COVID-19 to during COVID-19 for females or women versus males or men. Statistically significant, but small, sex- or gender-based differences were found in 2 of 8 mental health outcomes.

*Meaning:* Mental health changes among females or women were not significantly different from males or men for most outcomes, and differences that were identified were small and less than minimally important difference thresholds.

## INTRODUCTION

The COVID-19 pandemic has caused almost 6 million deaths and disrupted social and economic activities across the globe.^1,2^ By sex, males infected with COVID-19 are at greater risk of intensive care admission and death than females,^3,4^ but, by gender, socioeconomic burden has disproportionately impacted women.^5-12^ Economically, most single parents are women, and women earn less, are more likely to live in poverty, and hold less secure jobs than men, which heightens vulnerability.^5,8-11^ Women are overrepresented in health care jobs, which involves infection risk^5-10^ and provide most childcare and family elder care.^5,8-10^ Intimate partner violence has increased with the majority directed towards women.^5,7-10,12^ In addition to women, sex and gender minority individuals may face heightened socioeconomic challenges during COVID-19.^13,14^

Many of the socioeconomic implications of the pandemic that disproportionately affected women are known to be associated with worse mental health, and there is concern that increased burden during COVID-19 on women and gender minorities may have translated into worse mental health outcomes for these groups.^15-17^ Some researchers and prominent news media stories have reported that COVID-19 mental health effects have been greater for women than men.^18-28^ These reports, however, have been cross-sectional studies that evaluated proportions of participants above cut-offs on self-report measures without consideration of pre-COVID-19 differences, even though mental health disorders and symptoms were more common among women prior to the pandemic.^29-33^

Evidence from longitudinal cohorts that compare mental health symptoms pre-COVID-19 to during COVID-19 is needed to determine if there are gender differences. We are conducting a series of living systematic reviews on COVID-19 mental health,^34-36^ including mental health changes.^36^ The objective of this study was to compare mental health changes by sex or gender. This study goes beyond analyses presented in our main living systematic review, which reports on symptom levels and changes in many different population groups by conducting direct comparisons in the subset of studies that provide data on mental health changes by sex or gender.

## METHODS

Our series of living systematic reviews was registered in PROSPERO (CRD42020179703) and a protocol was posted to the Open Science Framework prior to initiating searches (https://osf.io/96csg/). The present study is a sub-study of our main mental health changes review.^36^ Results are reported per the PRISMA statement.^37^

### Study Eligibility

For our main symptom changes review, studies on any population were included if they compared mental health outcomes assessed between January 1, 2018 and December 31, 2019, when China first reported COVID-19 to the World Health Organization,^38^ to outcomes collected January 1, 2020 or later. We only included pre-COVID-19 data collected in the two years prior to COVID-19 to reduce comparisons of COVID-19 results with those collected during different developmental life stages. Compared samples had to include at least 90% of the same participants pre-COVID-19 and during COVID-19 or use statistical methods to account for missing follow-up data. Studies with < 100 participants were excluded for feasibility and due to their limited relative value. For the present analysis, studies had to report mental health outcomes separately by sex (assignment based on external genitalia, usually at birth; e.g., female, male, intersex) or gender (socially constructed characteristics of roles and behaviours; e.g., woman, man, trans woman, trans man, non-binary).^39^

### Search Strategy

MEDLINE (Ovid), PsycINFO (Ovid), CINAHL (EBSCO), EMBASE (Ovid), Web of Science Core Collection: Citation Indexes, China National Knowledge Infrastructure, Wanfang, medRxiv (preprints), and Open Science Framework Preprints (preprint server aggregator) were searched using a strategy designed by an experienced health science librarian. The China National Knowledge Infrastructure and Wanfang databases were searched using Chinese terms based on our English-language strategy. The rapid project launch did not allow for formal peer review, but COVID-19 terms were developed in collaboration with other librarians working on the topic. See Appendix 1 for search strategies. The initial search was conducted from December 31, 2019 to April 13, 2020 with automated daily updates. We converted to weekly updates on December 28, 2020 to increase processing efficiency.

### Selection of Eligible Studies

Search results were uploaded into DistillerSR (Evidence Partners, Ottawa, Canada). Duplicate references were removed. Then two reviewers independently evaluated titles and abstracts in random order; if either reviewer believed a study was potentially eligible, it underwent full-text review by two independent reviewers. Discrepancies at the full-text level were resolved by consensus, with a third reviewer consulted if necessary. An inclusion and exclusion coding guide was developed, and team members were trained over several sessions. See Appendix 2.

### Data Extraction

For each eligible study, data were extracted in DistillerSR by a single reviewer using a pre-specified form with validation by a second reviewer. Reviewers extracted (1) publication characteristics (e.g., first author, year, journal); (2) population characteristics and demographics, including eligibility criteria, recruitment method, number of participants, assessment timing, age; (3) mental health outcomes which included symptoms of anxiety, symptoms of depression, general mental health, and stress; (4) if studies reported outcomes by sex or gender or used these terms inconsistently (e.g., described using gender but reported results for females and males, which are sex terms); and (5) if sex or gender were treated as binary or categorical.

Adequacy of study methods and reporting was assessed using an adapted version of the Joanna Briggs Institute Checklist for Prevalence Studies, which assesses appropriateness of the sampling frame for the target population, appropriateness of recruiting methods, sample size, description of setting and participants, participation or response rate, outcome assessment methods, standardization of assessments across participants, appropriateness of statistical analyses, and follow-up.^40^ Each of the 9 items was coded as “yes” for meeting adequacy criteria, “no” for not meeting criteria, or “unclear” if incomplete reporting did not allow a judgment to be made. See Appendix 3.

For all data extraction, including adequacy of study methods and reporting, discrepancies were resolved between reviewers with a third reviewer consulted if necessary.

### Statistical Analyses

For continuous outcomes, separately for each sex or gender group, we extracted a standardized mean difference (SMD) effect size with 95% confidence intervals (CIs) for change from pre-COVID-19 to COVID-19. If not provided, we extracted pre-COVID-19 and COVID-19 means and standard deviations (SDs) for each group, calculated raw change scores (SD), and calculated SMD for change using Hedges’ g for each group,^41^ as described by Borenstein et al.^42^ Raw change scores were presented in scale units and direction, whereas SMD change scores were presented as positive when mental health worsened from pre-COVID-19 to COVID-19 and negative when it improved. We then calculated a Hedges’ g difference in change between sex or gender groups with 95% CI. Positive numbers represented greater negative change in females or women compared to males or men.

For studies that reported proportions of participants above a scale cut-off, for pre-COVID-19 and COVID-19 proportions, if not provided, we calculated a 95% CI using Agresti and Coull’s approximate method for binomial proportions.^43^ We then extracted or calculated the proportion change in participants above the cut-off, along with 95% CI, for each sex or gender group. Proportion changes were presented as positive when mental health worsened from pre-COVID-19 to COVID-19 and negative when it improved. If 95% CIs were not reported, we generated them using Newcombe’s method for differences between binomial proportions based on paired data.^44^ To do this, which requires the number of cases at both assessments, which is not typically available, we assumed that 50% of pre-COVID-19 cases continued to be cases during COVID-19 and confirmed that results did not differ substantively if we used values from 30% to 70% (all 95% CI end points within 0.02; see Appendix 4). Finally, we calculated a difference of the proportion change between sex or gender groups with 95% CI.^45^ Positive numbers reflected greater negative change in females or women compared to males or men.

Meta-analyses were done to synthesize differences between sex or gender groups in SMD change for continuous outcomes and in proportion change for dichotomous outcomes via restricted maximum-likelihood random-effects meta-analysis. Heterogeneity was assessed with the I^2^ statistic. Meta-analysis was performed in R (R version 3.6.3, RStudio Version 1.2.5042), using the metacont and metagen functions in the meta package.^46^ Forest plots were generated using the forest function in meta. Positive values indicated more relatively worse changes in mental health for females or women compared to males or men.

## RESULTS

### Search Results and Selection of Eligible Studies

As of August 30, 2021, there were 64,496 unique references identified and screened for potential eligibility, of which 63,534 were excluded after title and abstract review and 741 after full-text review. Of 221 remaining articles, 209 were excluded, leaving 12 included studies that reported data from 10 cohorts. Appendix 5 provides a figure that shows the flow of article review and reasons for exclusion.

### Characteristics of Included Studies

Four publications^47-50^ reported on 2 large, national, probability-based samples from the United Kingdom (N = 10,918 to 15,376)^47,48^ and the Netherlands (N = 3,983 to 4,064),^49,50^ and one publication^51^ reported on a community sample from Spain (N = 102). Two studies^52,53^ assessed young adults; one reported on a sample of twins from the United Kingdom (N = 3,563 to 3,694 depending on outcome)^52^ and another on a sample from Switzerland (N = 786).^53^ One study assessed adolescents from Australia (N = 248),^54^ and 3 studies^55-57^ assessed undergraduate students from China (N = 4,085 to 4,341),^55^ India (N = 217),^56^ and the United Kingdom (N = 214).^57^ One study^58^ assessed patients with systemic lupus erythematosus (N = 316). Four studies assessed anxiety symptoms,^52,54,56,58^ 4 depression symptoms,^52,54,56,58^ 7 (5 cohorts) general mental health,^47-51,55,57^ and 4 stress.^53,56-58^ Table 1 shows study characteristics. All studies compared women and men or females and males; none included other sex or gender groups. Use of sex and gender terms, however, was inconsistent in 5 of 12 included studies^48,52,54,56,57^ (e.g., described assessing gender but reporting results for “females” and “males”). Results during COVID-19 were assessed between March and June 2020 for all studies. Two cohorts also reported results from September 2020^48^ and November to December 2020.^50^

**Table 1.**
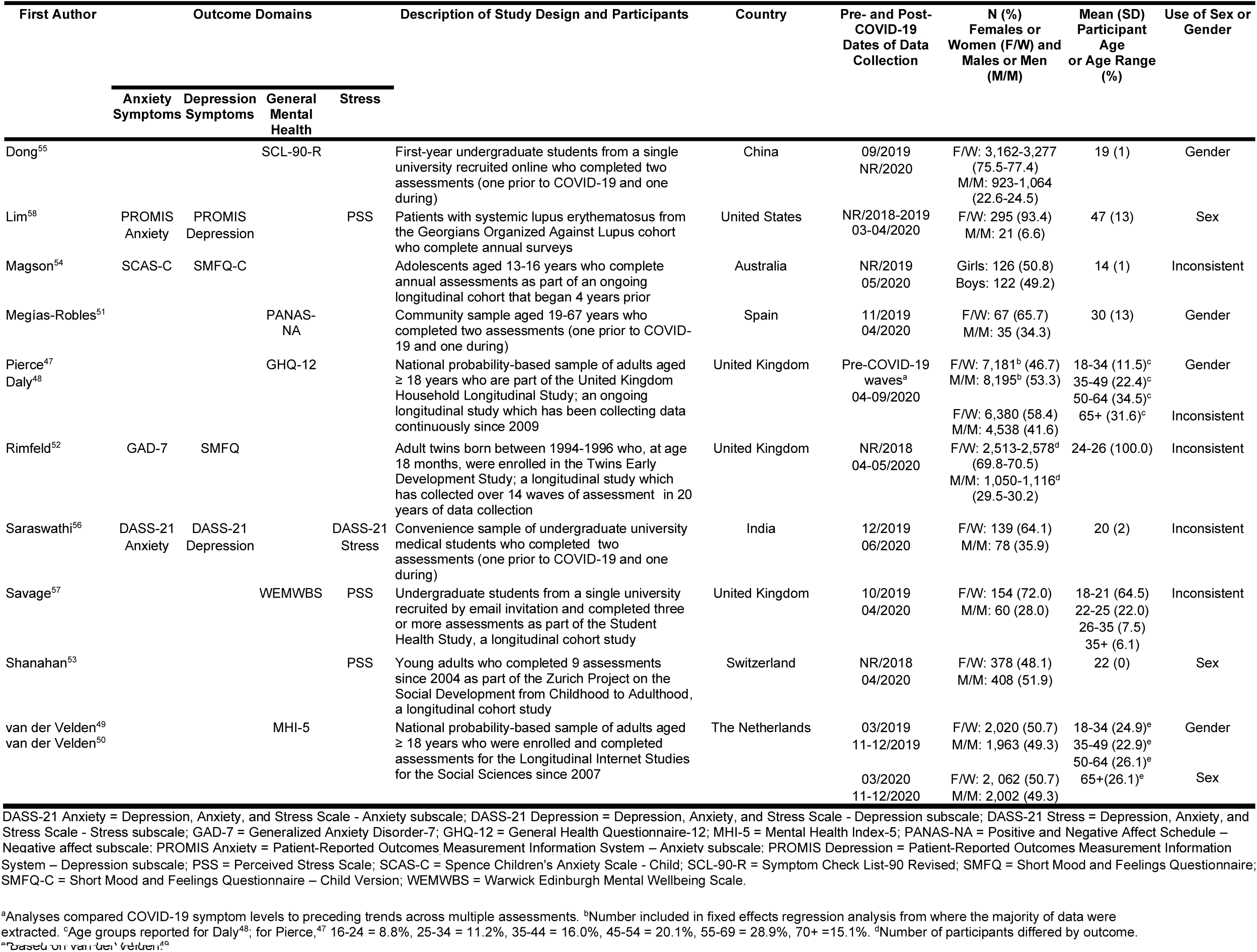
Characteristics of Included Studies (N=12)

### Adequacy of Study Methods and Reporting

Two studies (1 cohort)^49,50^ were rated as “yes” for adequacy for all items. Other studies were rated “no” for 1-3 items (plus 0-3 unclear ratings)^48,51,53-57^ or “no” on none but unclear on 2-4 items.^47,52,58^ There were 6 studies^51,53-57^ rated “no” or “unclear” for appropriate sampling frame (50.0%), 8 “no” or “unclear” for adequate response rate and coverage (66.7%),^47,48,51-54,57,58^ and 7 “no” or “unclear” for follow-up response rate and management (58.3%).^47,48,51,52,54,57,58^ See Appendix 6 for results for all studies.

### Mental Health Symptom Changes

There was a total of 15 comparisons of continuous score changes and 6 of proportion changes; in 15 out of 21 comparisons, females or women had worse mental health pre-COVID-19. Mental health scores and symptom changes for all outcome domains are reported separately by sex or gender groups in Table 2. Differences in continuous and dichotomous changes by sex or gender are shown in Figures 1 and 2. Estimates of difference in change by sex or gender were close to zero and not statistically significant for anxiety symptoms with dichotomous outcomes (Figure 2a; proportion change difference = -0.05, 95% CI -0.20 to 0.11; N = 1 study,^56^ 217 participants), depression symptoms with continuous (Figure 1b; SMD change difference = 0.12, 95% CI -0.09 to 0.33; N = 4 studies,^52,54,56,58^ 4,475 participants; I^2^ = 69.0%) and dichotomous outcomes (Figure 2b; proportion change difference = 0.12, 95% CI -0.03 to 0.28; N = 1 study,^56^ 217 participants), general mental health dichotomous outcomes (Figure 2c [all results from early 2020]; proportion change difference = -0.03, 95% CI -0.09 to 0.04; N = 3 studies,^48,49,55^ 18,985 participants; I^2^ = 94.0%), and stress with continuous (Figure 1d; SMD change difference = -0.10, 95% CI -0.21 to 0.01; N = 4 studies,^53,56-58^ 1,533 participants; I^2^ = 0.0%) and dichotomous outcomes (Figure 2d; proportion change difference = 0.04, 95% CI - 0.10 to 0.17; N = 1 study,^56^ 217 participants). Of the 4 studies^48-50,55^ that reported dichotomous general mental health, 2 studies^48,50^ also reported outcomes from late 2020; when those results were used, the null finding did not change (Figure 2e; proportion change difference = 0.00, 95% CI -0.03 to 0.03; N = 3 studies,^48,50,55^ 19,067 participants; I^2^ = 67.0%).

**Table 2.**
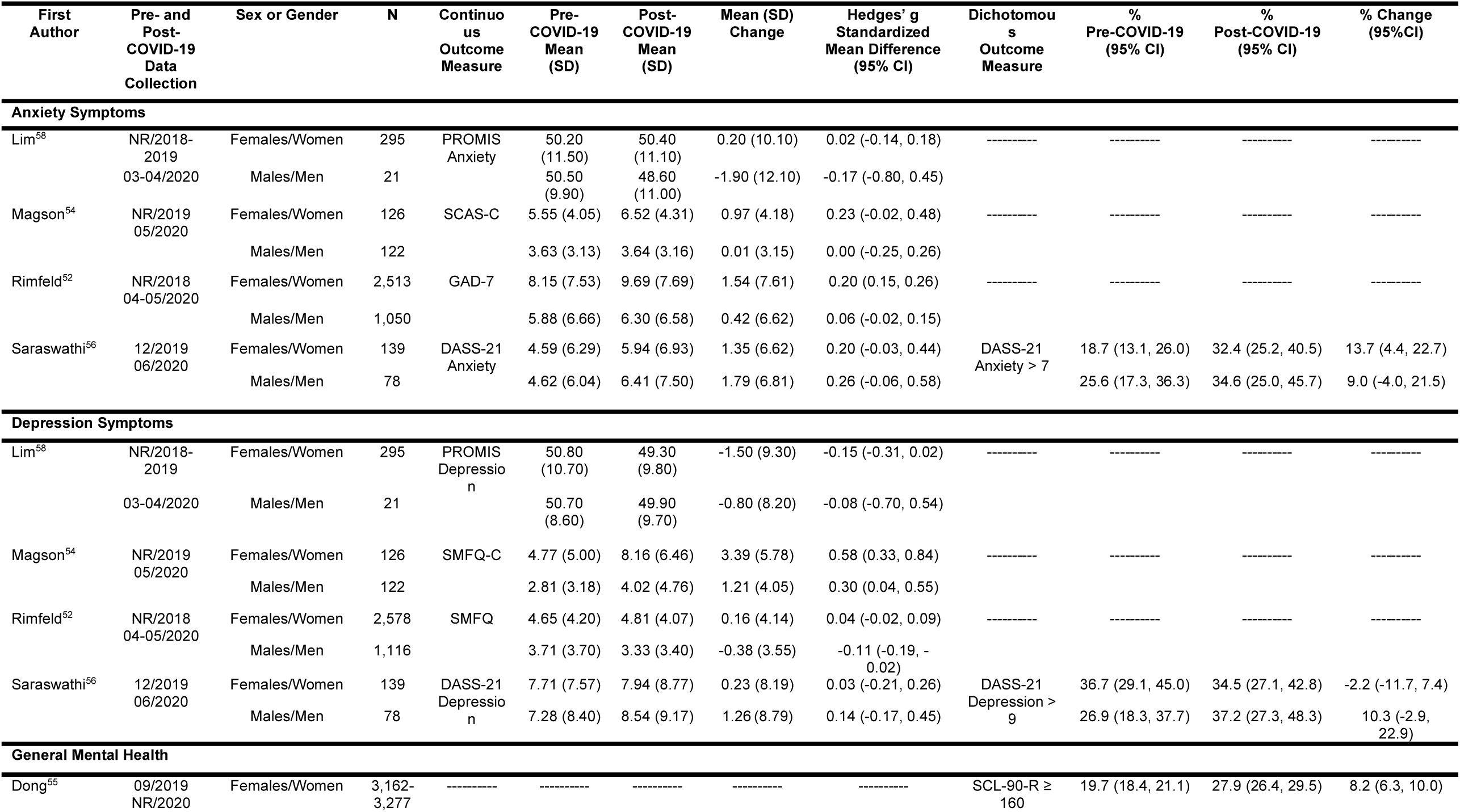
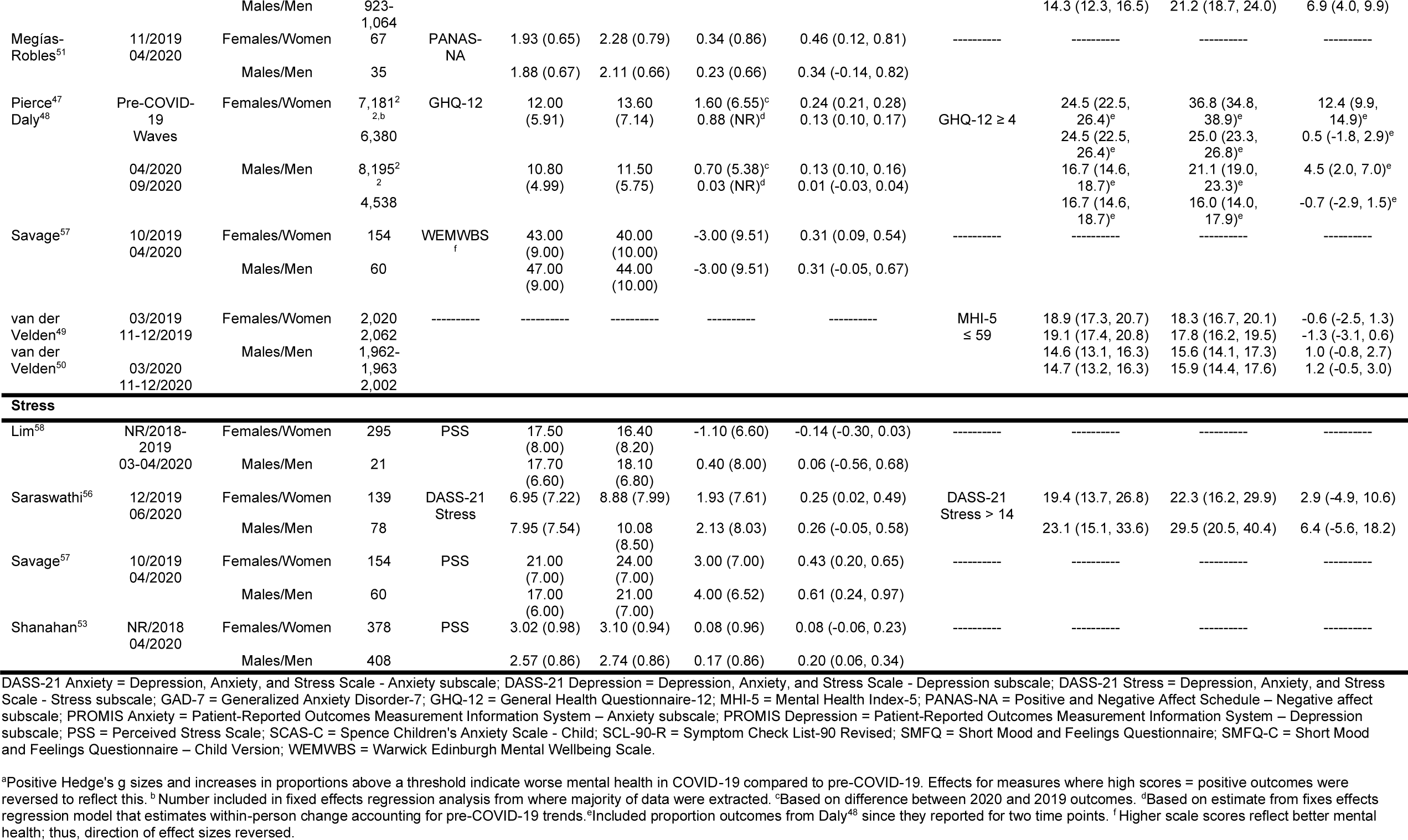
Outcomes From Included Studies by Sex or Gender^a^.

| First Author | Pre- and Post-COVID-19 Data Collection | Sex or Gender | N | Continuous Outcome Measure | Pre-COVID-19 Mean (SD) | Post-COVID-19 Mean (SD) | Mean (SD) Change | Hedges' g Standardized Mean Difference (95% CI) | Dichotomous Outcome Measure | % Pre-COVID-19 (95% CI) | % Post-COVID-19 (95% CI) | % Change (95%CI) |
| --- | --- | --- | --- | --- | --- | --- | --- | --- | --- | --- | --- | --- |
| Anxiety Symptoms |  |  |  |  |  |  |  |  |  |  |  |  |
| Lim<sup>58</sup> | NR/2018-2019 | Females/Women | 295 | PROMIS Anxiety | 50.20 (11.50) | 50.40 (11.10) | 0.20 (10.10) | 0.02 (-0.14, 0.18) | ----- | ----- | ----- | ----- |
|  | 03-04/2020 | Males/Men | 21 |  | 50.50 (9.90) | 48.60 (11.00) | -1.90 (12.10) | -0.17 (-0.80, 0.45) |  |  |  |  |
| Magson<sup>54</sup> | NR/2019 05/2020 | Females/Women | 126 | SCAS-C | 5.55 (4.05) | 6.52 (4.31) | 0.97 (4.18) | 0.23 (-0.02, 0.48) | ----- | ----- | ----- | ----- |
|  |  | Males/Men | 122 |  | 3.63 (3.13) | 3.64 (3.16) | 0.01 (3.15) | 0.00 (-0.25, 0.26) |  |  |  |  |
| Rimfeld<sup>52</sup> | NR/2018 04-05/2020 | Females/Women | 2,513 | GAD-7 | 8.15 (7.53) | 9.69 (7.69) | 1.54 (7.61) | 0.20 (0.15, 0.26) | ----- | ----- | ----- | ----- |
|  |  | Males/Men | 1,050 |  | 5.88 (6.66) | 6.30 (6.58) | 0.42 (6.62) | 0.06 (-0.02, 0.15) |  |  |  |  |
| Saraswathi<sup>56</sup> | 12/2019 06/2020 | Females/Women | 139 | DASS-21 Anxiety | 4.59 (6.29) | 5.94 (6.93) | 1.35 (6.62) | 0.20 (-0.03, 0.44) | DASS-21 Anxiety > 7 | 18.7 (13.1, 26.0) | 32.4 (25.2, 40.5) | 13.7 (4.4, 22.7) |
|  |  | Males/Men | 78 |  | 4.62 (6.04) | 6.41 (7.50) | 1.79 (6.81) | 0.26 (-0.06, 0.58) |  | 25.6 (17.3, 36.3) | 34.6 (25.0, 45.7) | 9.0 (-4.0, 21.5) |
| Depression Symptoms |  |  |  |  |  |  |  |  |  |  |  |  |
| Lim<sup>58</sup> | NR/2018-2019 | Females/Women | 295 | PROMIS Depression | 50.80 (10.70) | 49.30 (9.80) | -1.50 (9.30) | -0.15 (-0.31, 0.02) | ----- | ----- | ----- | ----- |
|  | 03-04/2020 | Males/Men | 21 |  | 50.70 (8.60) | 49.90 (9.70) | -0.80 (8.20) | -0.08 (-0.70, 0.54) |  |  |  |  |
| Magson<sup>54</sup> | NR/2019 05/2020 | Females/Women | 126 | SMFQ-C | 4.77 (5.00) | 8.16 (6.46) | 3.39 (5.78) | 0.58 (0.33, 0.84) | ----- | ----- | ----- | ----- |
|  |  | Males/Men | 122 |  | 2.81 (3.18) | 4.02 (4.76) | 1.21 (4.05) | 0.30 (0.04, 0.55) |  |  |  |  |
| Rimfeld<sup>52</sup> | NR/2018 04-05/2020 | Females/Women | 2,578 | SMFQ | 4.65 (4.20) | 4.81 (4.07) | 0.16 (4.14) | 0.04 (-0.02, 0.09) | ----- | ----- | ----- | ----- |
|  |  | Males/Men | 1,116 |  | 3.71 (3.70) | 3.33 (3.40) | -0.38 (3.55) | -0.11 (-0.19, -0.02) |  |  |  |  |
| Saraswathi<sup>56</sup> | 12/2019 06/2020 | Females/Women | 139 | DASS-21 Depression | 7.71 (7.57) | 7.94 (8.77) | 0.23 (8.19) | 0.03 (-0.21, 0.26) | DASS-21 Depression > 9 | 36.7 (29.1, 45.0) | 34.5 (27.1, 42.8) | -2.2 (-11.7, 7.4) |
|  |  | Males/Men | 78 |  | 7.28 (8.40) | 8.54 (9.17) | 1.26 (8.79) | 0.14 (-0.17, 0.45) |  | 26.9 (18.3, 37.7) | 37.2 (27.3, 48.3) | 10.3 (-2.9, 22.9) |
| General Mental Health |  |  |  |  |  |  |  |  |  |  |  |  |
| Dong<sup>55</sup> | 09/2019 NR/2020 | Females/Women | 3,162-3,277 | ----- | ----- | ----- | ----- | ----- | SCL-90-R ≥ 160 | 19.7 (18.4, 21.1) | 27.9 (26.4, 29.5) | 8.2 (6.3, 10.0) |
|  |  | Males/Men | 923-1,064 |  |  |  |  |  |  | 14.3 (12.3, 16.5) | 21.2 (18.7, 24.0) | 6.9 (4.0, 9.9) |
| Megías-Robles<sup>51</sup> | 11/2019<br>04/2020 | Females/Women | 67 | PANAS-NA | 1.93 (0.65) | 2.28 (0.79) | 0.34 (0.86) | 0.46 (0.12, 0.81) | ----- | ----- | ----- | ----- |
|  |  | Males/Men | 35 |  | 1.88 (0.67) | 2.11 (0.66) | 0.23 (0.66) | 0.34 (-0.14, 0.82) |  |  |  |  |
| Pierce<sup>47</sup><br>Daly<sup>48</sup> | Pre-COVID-19<br>Waves | Females/Women | 7,181<sup>2</sup><br><sub>2,b</sub><br>6,380 | GHQ-12 | 12.00<br>(5.91) | 13.60<br>(7.14) | 1.60 (6.55)<sup>c</sup><br>0.88 (NR)<sup>d</sup> | 0.24 (0.21, 0.28)<br>0.13 (0.10, 0.17) | GHQ-12 ≥ 4 | 24.5 (22.5, 26.4)<sup>e</sup><br>24.5 (22.5, 26.4)<sup>e</sup> | 36.8 (34.8, 38.9)<sup>e</sup><br>25.0 (23.3, 26.8)<sup>e</sup> | 12.4 (9.9, 14.9)<sup>e</sup><br>0.5 (-1.8, 2.9)<sup>e</sup> |
|  | 04/2020<br>09/2020 | Males/Men | 8,195<sup>2</sup><br><sub>2</sub><br>4,538 |  | 10.80<br>(4.99) | 11.50<br>(5.75) | 0.70 (5.38)<sup>c</sup><br>0.03 (NR)<sup>d</sup> | 0.13 (0.10, 0.16)<br>0.01 (-0.03, 0.04) |  | 16.7 (14.6, 18.7)<sup>e</sup><br>16.7 (14.6, 18.7)<sup>e</sup> | 21.1 (19.0, 23.3)<sup>e</sup><br>16.0 (14.0, 17.9)<sup>e</sup> | 4.5 (2.0, 7.0)<sup>e</sup><br>-0.7 (-2.9, 1.5)<sup>e</sup> |
| Savage<sup>57</sup> | 10/2019<br>04/2020 | Females/Women | 154 | WEMWBS<sup>f</sup> | 43.00<br>(9.00) | 40.00<br>(10.00) | -3.00 (9.51) | 0.31 (0.09, 0.54) | ----- | ----- | ----- | ----- |
|  |  | Males/Men | 60 |  | 47.00<br>(9.00) | 44.00<br>(10.00) | -3.00 (9.51) | 0.31 (-0.05, 0.67) |  |  |  |  |
| van der Velden<sup>49</sup> | 03/2019<br>11-12/2019 | Females/Women | 2,020<br>2,062 | ----- | ----- | ----- | ----- | ----- | MHI-5 ≤ 59 | 18.9 (17.3, 20.7)<br>19.1 (17.4, 20.8) | 18.3 (16.7, 20.1)<br>17.8 (16.2, 19.5) | -0.6 (-2.5, 1.3)<br>-1.3 (-3.1, 0.6) |
| van der Velden<sup>50</sup> | 03/2020<br>11-12/2020 | Males/Men | 1,962-1,963<br>2,002 |  |  |  |  |  |  | 14.6 (13.1, 16.3)<br>14.7 (13.2, 16.3) | 15.6 (14.1, 17.3)<br>15.9 (14.4, 17.6) | 1.0 (-0.8, 2.7)<br>1.2 (-0.5, 3.0) |
| **Stress** |  |  |  |  |  |  |  |  |  |  |  |  |
| Lim<sup>58</sup> | NR/2018-2019<br>03-04/2020 | Females/Women | 295 | PSS | 17.50<br>(8.00) | 16.40<br>(8.20) | -1.10 (6.60) | -0.14 (-0.30, 0.03) | ----- | ----- | ----- | ----- |
|  |  | Males/Men | 21 |  | 17.70<br>(6.60) | 18.10<br>(6.80) | 0.40 (8.00) | 0.06 (-0.56, 0.68) |  |  |  |  |
| Saraswathi<sup>56</sup> | 12/2019<br>06/2020 | Females/Women | 139 | DASS-21 Stress | 6.95 (7.22) | 8.88 (7.99) | 1.93 (7.61) | 0.25 (0.02, 0.49) | DASS-21 Stress > 14 | 19.4 (13.7, 26.8) | 22.3 (16.2, 29.9) | 2.9 (-4.9, 10.6) |
|  |  | Males/Men | 78 |  | 7.95 (7.54) | 10.08<br>(8.50) | 2.13 (8.03) | 0.26 (-0.05, 0.58) |  | 23.1 (15.1, 33.6) | 29.5 (20.5, 40.4) | 6.4 (-5.6, 18.2) |
| Savage<sup>57</sup> | 10/2019<br>04/2020 | Females/Women | 154 | PSS | 21.00<br>(7.00) | 24.00<br>(7.00) | 3.00 (7.00) | 0.43 (0.20, 0.65) | ----- | ----- | ----- | ----- |
|  |  | Males/Men | 60 |  | 17.00<br>(6.00) | 21.00<br>(7.00) | 4.00 (6.52) | 0.61 (0.24, 0.97) |  |  |  |  |
| Shanahan<sup>53</sup> | NR/2018<br>04/2020 | Females/Women | 378 | PSS | 3.02 (0.98) | 3.10 (0.94) | 0.08 (0.96) | 0.08 (-0.06, 0.23) | ----- | ----- | ----- | ----- |
|  |  | Males/Men | 408 |  | 2.57 (0.86) | 2.74 (0.86) | 0.17 (0.86) | 0.20 (0.06, 0.34) |  |  |  |  |

DASS-21 Anxiety = Depression, Anxiety, and Stress Scale - Anxiety subscale; DASS-21 Depression = Depression, Anxiety, and Stress Scale - Depression subscale; DASS-21 Stress = Depression, Anxiety, and Stress Scale - Stress subscale; GAD-7 = Generalized Anxiety Disorder-7; GHQ-12 = General Health Questionnaire-12; MHI-5 = Mental Health Index-5; PANAS-NA = Positive and Negative Affect Schedule – Negative affect subscale; PROMIS Anxiety = Patient-Reported Outcomes Measurement Information System – Anxiety subscale; PROMIS Depression = Patient-Reported Outcomes Measurement Information System – Depression subscale; PSS = Perceived Stress Scale; SCAS-C = Spence Children's Anxiety Scale - Child; SCL-90-R = Symptom Check List-90 Revised; SMFQ = Short Mood and Feelings Questionnaire; SMFQ-C = Short Mood and Feelings Questionnaire – Child Version; WEMWBS = Warwick Edinburgh Mental Wellbeing Scale.
<sup>a</sup>Positive Hedge's g sizes and increases in proportions above a threshold indicate worse mental health in COVID-19 compared to pre-COVID-19. Effects for measures where high scores = positive outcomes were reversed to reflect this. <sup>b</sup>Number included in fixed effects regression analysis from where majority of data were extracted. <sup>c</sup>Based on difference between 2020 and 2019 outcomes. <sup>d</sup>Based on estimate from fixed effects regression model that estimates within-person change accounting for pre-COVID-19 trends. <sup>e</sup>Included proportion outcomes from Daly<sup>48</sup> since they reported for two time points. <sup>f</sup>Higher scale scores reflect better mental health; thus, direction of effect sizes reversed.

**Figure 1a-1d.**
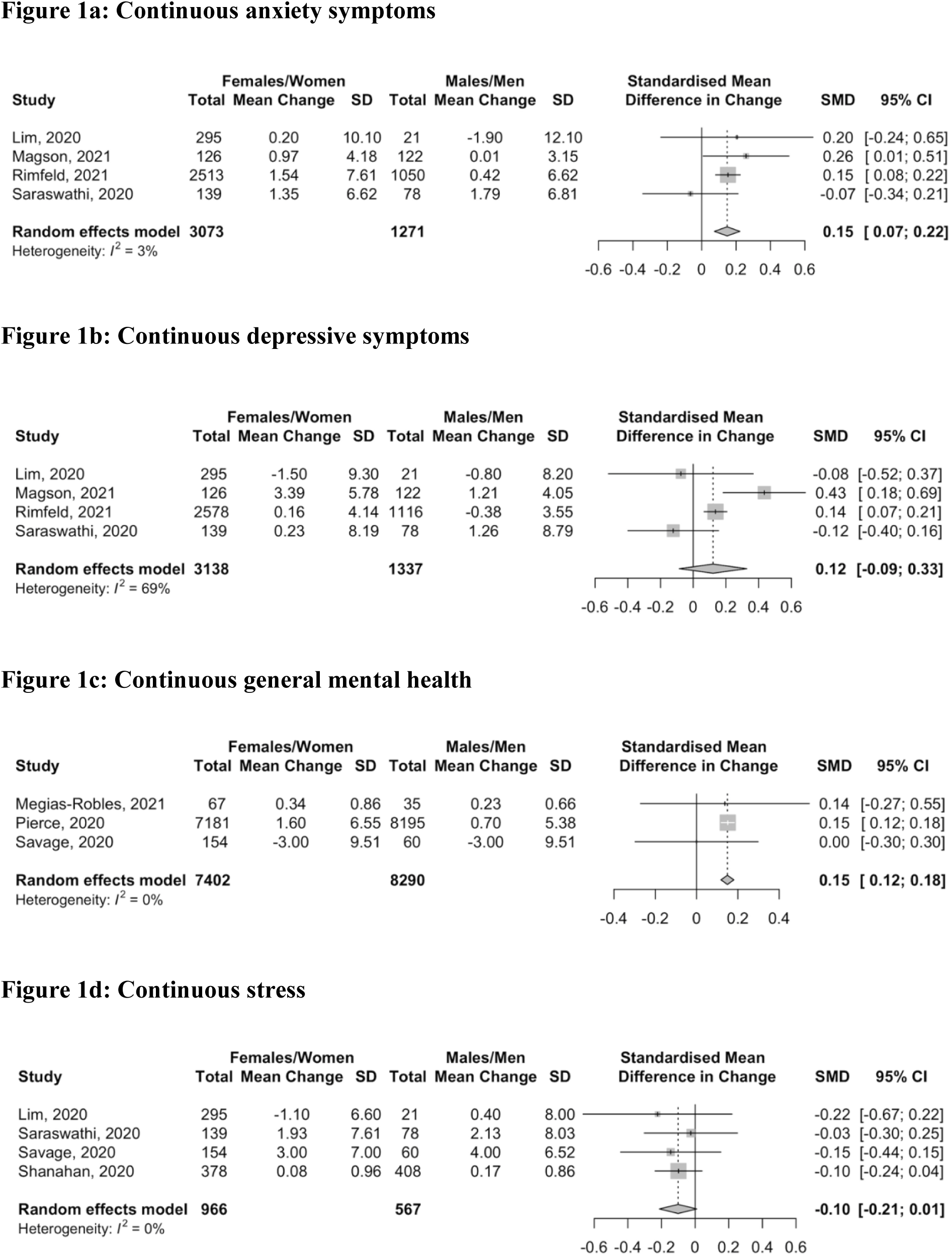
Forest plots of standardized mean difference of the difference in change in continuous anxiety symptom scores (1a), depression symptom scores (1b), general mental health scores (1c), and stress scores (1d) between females or women and males or men. Positive numbers indicate greater negative change in mental health in females or women compared to males or men.

**Figure 2a-2e.**
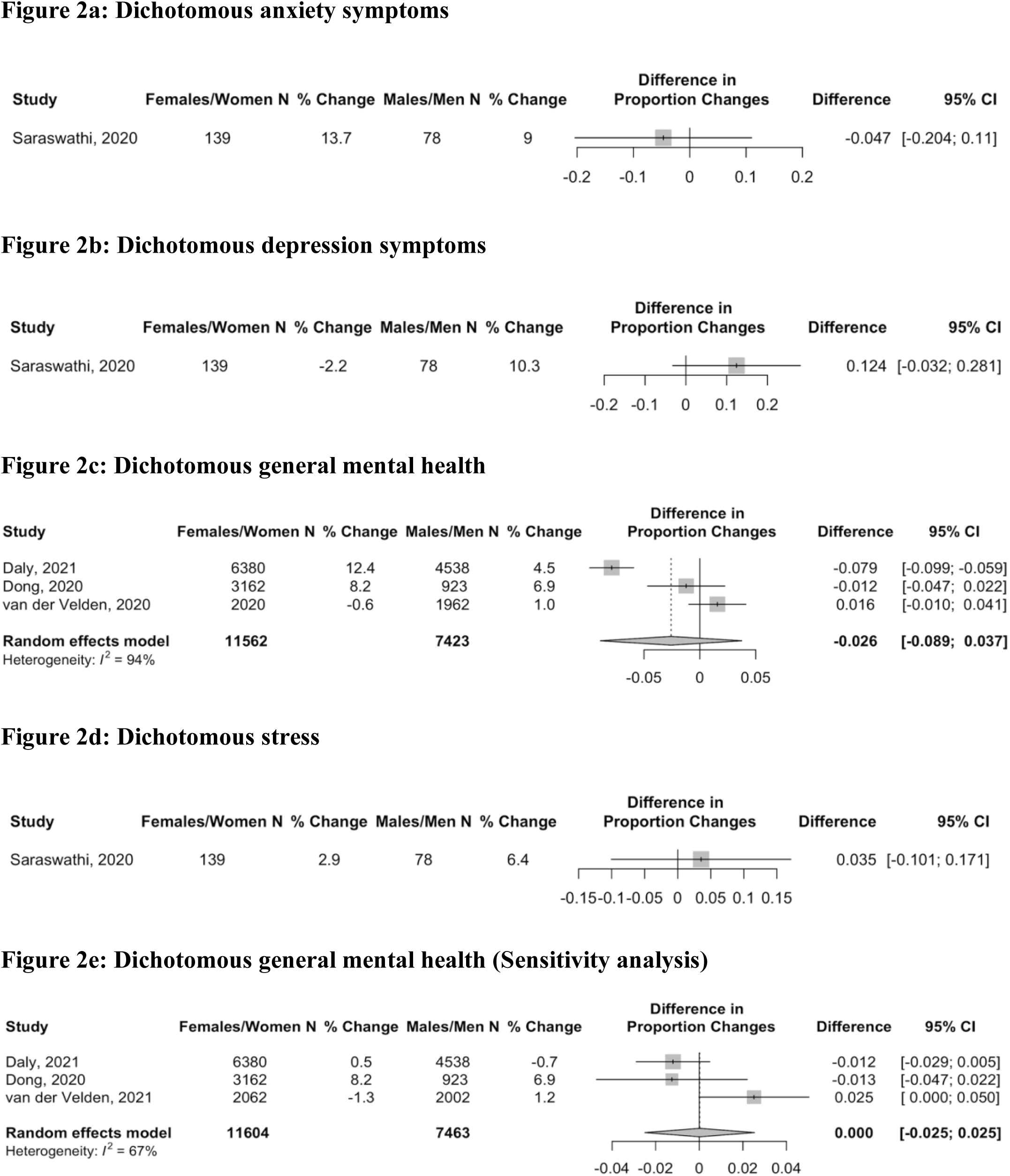
Forest plots of standardized mean difference of the difference in change in proportion above a cut-off for anxiety (2a), depression (2b), general mental health (2c), and stress (2d) between females or women and males or men. Positive numbers indicate greater negative change in mental health in females or women compared to males or men. Figure 2c reflects dichotomous COVID-19 mental health measured in early 2020, whereas 2e reflects measurements from late 2020 for Daly^48^ and van der Velden.^50^

Anxiety, measured continuously, worsened significantly more for females or women than for males or men during COVID-19 (Figure 1a; SMD change difference = 0.15, 95% CI 0.07 to 0.22; N = 4 studies,^52,54,56,58^ 4,344 participants; I^2^= 3.0%). General mental health, measured continuously, also worsened more for females or women than for males or men in early COVID-19 (Figure 1c; SMD difference in change = 0.15, 95% CI 0.12 to 0.18; N = 3 studies,^47,51,57^ 15,692 participants; I^2^ = 0.0%). This was predominantly based on a large population-based study from the United Kingdom.^47^ That study did not report results from fall 2020 for continuous outcomes, but as shown in Table 2 and Figures 2c and 2e, the difference in change between females or women and males or men decreased between early and late 2020 for dichotomous outcomes in the same cohort.^48^ The magnitude of both statistically significant differences was small (see Figure 3).

**Figure 3.**
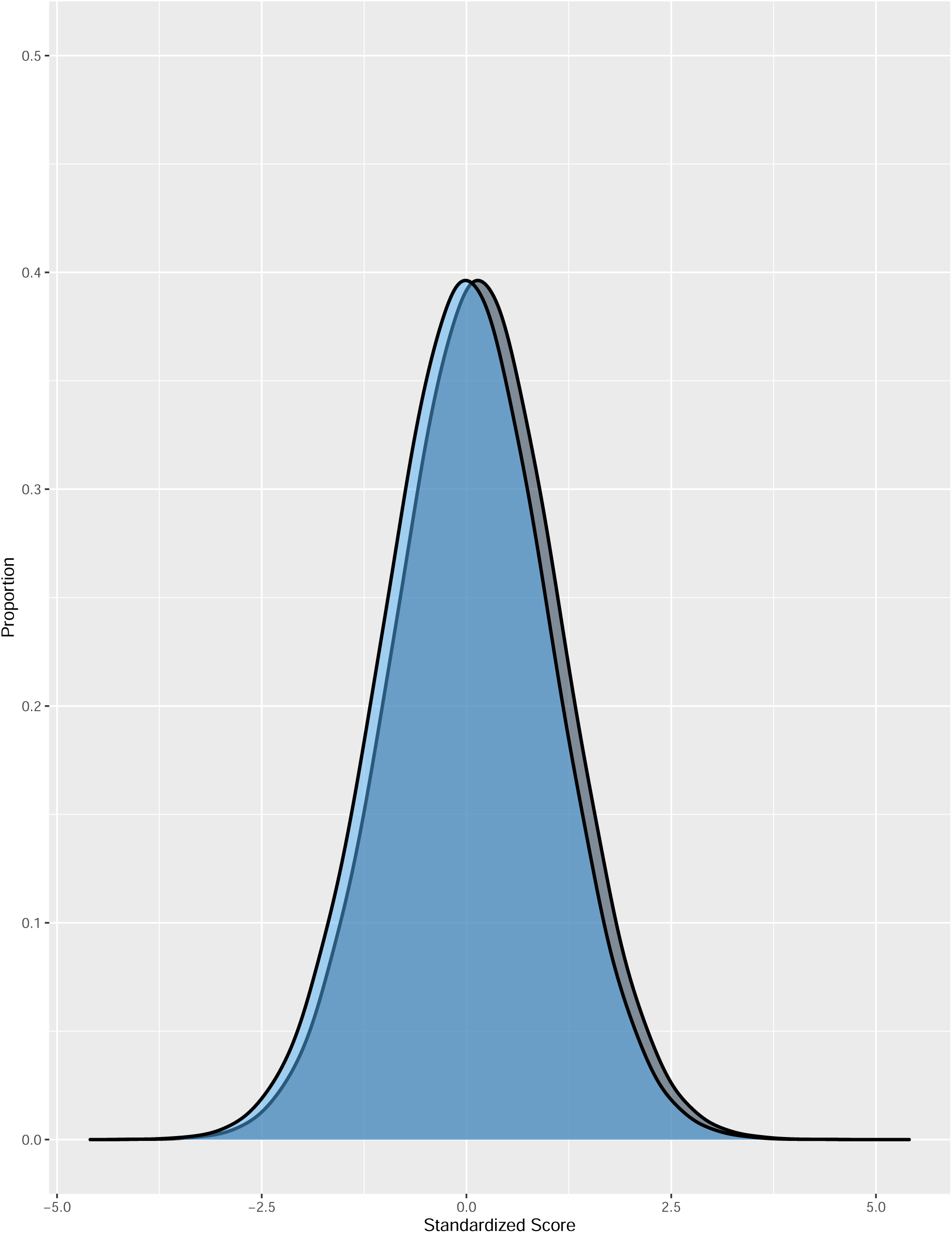
Illustration of the magnitude of change for SMD = 0.15 assuming a normal distribution. The hypothetical blue distribution represents pre-COVID-19 scores, and the grey distribution represents post-COVID-19 scores with a mean symptom increase of SMD = 0.15.

## DISCUSSION

The COVID-19 pandemic has affected women and gender minorities disproportionately.^5-14^ There has been an assumption, seemingly confirmed by cross-sectional data collected during COVID-19, that this has translated into greater negative changes in mental health among women.^18-28^ We reviewed evidence from 12 studies (10 cohorts) that reported mental health changes from pre-COVID-19 to COVID-19 separately by sex or gender. We compared females or women with males or men; no studies compared gender minorities with any other group. Syntheses of continuously measured anxiety symptoms (SMD = 0.15, 95% CI 0.07 to 0.22) and general mental health (SMD = 0.15, 95% CI 0.12 to 0.18) found that mental health worsened more for females or women than males or men, but the magnitude was small and far below thresholds that are typically considered clinically important (e.g., SMD = 0.50).^59^ None of the other 6 mental health outcomes that we examined (continuous depression symptoms and stress; dichotomous anxiety symptoms, depression symptoms, general mental health, and stress) differed by sex or gender.

Sex and gender differences in mental health disorder prevalence, symptoms, and risk factors are well-established.^60-63^ Likely risk factors include gender inequities and discrimination, economic disadvantage and poverty, higher rates of interpersonal stressors, and violence,^64,65^ and many of these risk factors have been exacerbated for women during COVID-19.^5-12^ We did not identify any differences in mental health by sex or gender, however, that appeared to be substantive; all were 0.15 SMD or smaller, which is considered to be a small difference based on commonly used metrics (e.g., < 0.20 SMD)^66^ and below thresholds for clinical meaningfulness.^59^

Based on our findings, it is possible that despite the challenges women have faced, many have been resilient and that the mental health disaster that has been predicted by many has not occurred.^67^ This finding departs from what has been reported in some research and by the media. Three factors may feed this discrepancy. One is the publication of many cross-sectional studies that report proportions above cut-offs on self-report measures, which are not designed for that purpose,^68-72^ and assume that what are perceived as high numbers, generally, or sex differences, comparatively, must not have been present pre-COVID-19.^36^ A second is the use of surveys that ask questions about well-being with COVID-19 explicitly assigned as a cause; illustrating the pitfalls of this, a study of over 2,000 young Swiss adult men found significant angst when questions were asked in this way, but no changes in validated measures of depression symptoms and stress from pre-COVID.^73^ A third reason relates to news media reports that emphasize dramatic events and anecdotes without evidence that demonstrates changes.^67^

Strengths of our study include the use of rigorous systematic review methods. We searched 9 databases, including Chinese-language databases, without language restrictions and included studies that enabled the direct comparison of mental health changes by sex or gender. There are limitations to consider. First, this review only included 12 studies from 10 cohorts, and many had limitations related to study sampling frames and recruitment methods, follow-up rates, and management of missing data. Second, heterogeneity was high for some meta-analyses; it was low, however, for others, and results across 8 analyses did not differ substantively. Third, there were not enough studies to attempt sub-group analyses by additional sociodemographic or other factors. Fourth, we did not identify any studies that compared results from gender-diverse individuals to other gender groups. Fifth, in calculating 95% CIs for within-group changes in proportions with the information provided in publications (pre-COVID-19 and COVID-19 group proportions), we assumed that 50% of pre-COVID-19 cases continued to be cases during COVID-19. However, the maximum difference in any end point of a 95% CIs across analyses was 0.02 when we varied our assumption from 30% to 70%.

In sum, we identified small sex- or gender-based differences for anxiety symptoms and general mental health, continuously measured, but other outcomes (continuous depression symptoms and stress; dichotomous anxiety symptoms, depression symptoms, general mental health, and stress) did not differ by sex or gender. This finding diverges from what has been reported from cross-sectional studies. These are aggregate results, though, and many individuals have certainly experienced negative mental health changes related to increased socioeconomic burden. It seems plausible, given the divergent ways that the pandemic has affected different people that many people are experiencing improved mental health, whereas large numbers of others may be experiencing worsened mental health, including new onset mental disorders among people without previous morbidity. Thus, mental health changes should continue to be monitored in COVID-19, taking into consideration sex and gender, and studies should examine reasons for what appears to be resilience among many women despite facing disproportionate hardships in the pandemic. Health care providers should be alert to life changes that may be associated with vulnerability and to physical and emotional or cognitive symptoms that may reflect worsening mental health so that they can assess, if appropriate, and provide mental health care to those in need.

## Data Availability

All data used in the study are available in the manuscript and its tables or online at
https://www.depressd.ca/covid-19-mental-health.

## Author Contributions

YS, YWu, DBR, AB, and BDT were responsible for the study conception and design. JTB was responsible for the design of the database searches. AK carried out the searches. TDS, YS, YWu, CH, YWang, XJ, KL, OB, AK, DBR, SM, MA, DN, AT, AY, ITV and BDT contributed to data extraction, coding, and evaluation of included studies. YS, CH, and OB were responsible for study coordination. TDS, YS, YWu, BL, AB, and BDT were involved in data analysis. BA, CF, MSM, SS, and GT contributed to interpretation of results as knowledge translation partners. TDS and BDT drafted the manuscript. All authors provided a critical review and approved the final manuscript. BDT is the guarantor; he had full access to all the data in the study and takes responsibility for the integrity of the data and the accuracy of the data analyses. BDT is the corresponding author and attests that all listed authors meet authorship criteria and that no others meeting the criteria have been omitted.

## Data Sharing

All data used in the study are available in the manuscript and its tables or online at https://www.depressd.ca/covid-19-mental-health.

