## Supplemental Files for "Comparison of Mental Health Symptom Changes from pre-COVID-19 to COVID-19 by Sex or Gender: A Systematic Review and Meta-analysis"

**SUPPLEMENTARY FILES**

**Appendix 1.** Search Strategies

**Appendix 2.** Title and Abstract and Full-Text Review Inclusion and Exclusion Criteria Coding Guides

**Appendix 3.** Adequacy of Study Methods and Reporting

**Appendix 4.** Confidence Intervals for Proportion Change When Using 30% and 70% of Pre-COVID-19 Cases were Cases During COVID-19: Calculated for Studies that Did Not Provide 95% Confidence Intervals for Change

**Appendix 5.** PRISMA Flowchart

**Appendix 6.** Adequacy of Methods and Reporting of Included Studies

**Appendix 1. Search Strategies**

Search strategies can be found in the project folder on the Open Science Framework:

<https://osf.io/96csg/>

**Appendix 2. Title and Abstract and Full-Text Review Inclusion and Exclusion Criteria Coding Guides**

**Title and Abstract Coding Criteria**

**MENTAL HEALTH SYMPTOM CHANGES CODING CRITERIA:**

**No: not original human data or a case study or case series.** If it is clear from the title and abstract that the article is not an original report of primary data, but, for example, a letter, editorial, systematic review or meta-analysis, or it is a single case study or case series, then it is excluded. Studies reporting only on animal, cellular, or genetic data are also excluded. Conference abstracts are included.

**No: not a study of any population affected by the COVID-19 outbreak.** If it is clear from the title or abstract that the study is not about any population affected by the COVID-19 outbreak, it is excluded. Studies that include fewer than 100 subjects, are excluded. If a longitudinal study has baseline sample size with at least 100 participants, but no follow-up with at least 100 participants, then we exclude the study (and document); if its baseline and at least one follow-up have more than 100 participants, we include the study.

**No: not a study which reports mental health symptom changes longitudinally pre-COVID-19 to COVID-19 or during COVID-19.** If it is clear from the title or abstract that the study does not report continuous scores of symptom levels or proportions of participants meeting the threshold on a validated scale, or diagnostic criteria using a validated diagnostic interview prior to and after the start of COVID-19, or longitudinally during COVID-19, then it will be excluded.

For pre-COVID versus during-COVID studies, pre- and during- samples must include the same cohort, not different representative samples. Pre- and during-samples should have less than 10% difference in the participants in the sample* or should statistically account for missing data, i.e., if N between the samples differs by more than 10%, modelling or imputation is needed to evaluate results for all participants. Pre-COVID data needs to be collected prior to 2020 (or at least 80% of the participants’ data need be collected prior to 2020 if collection spans from 2019 to 2020) and after 2018 (or at least 80% of the participants’ data need to be collected after 2018 if collection spans from pre-2018 to 2018).

For studies with multiple waves across COVID, if there are pre-pandemic time points, the most recent pre-pandemic wave needs to be in 2018 or later; if the most recent pre-pandemic wave spans from pre-2018 to 2018, at least 80% of the data need to be collected in 2018. Studies with multiple waves across COVID-19 must have at least two time points that have less than 10% difference in the participants in the sample*, or should statistically account for missing data, regardless of whether or not the study has pre-COVID assessments.

If outcomes from the study are only shown graphically without eligible numerical values, exclude the study.

* At least 90% of participants in assessments from two time points need to be the same participants. In a three-wave survey, if N-T1 = 1000, N – T2 = 500, and N – T3 = 500, T2 and T3 would only be eligible if at least 90% of the participants at each time point were the same. It is not enough to just have a total N within 10%.

**Yes: study eligible for inclusion in full-text review.**

**Full Text Coding Criteria**

**MENTAL HEALTH SYMPTOM CHANGES CODING CRITERIA:**

**No: not original human data or a case study or case series.** If the article is not an original report of primary data, but, for example, a letter, editorial, systematic review or meta-analysis, or it is a single case study or case series, then it is excluded. Studies reporting only on animal, cellular, or genetic data are also excluded. Conference abstracts are included.

**No: not a study of any population affected by the COVID-19 outbreak.** If the study is not about any population affected by the COVID-19 outbreak, it is excluded. Studies that include fewer than 100 subjects, are excluded. If a longitudinal study has baseline sample size with at least 100 participants, but no follow-up with at least 100 participants, then we exclude the study (and document); if its baseline and at least one follow-up have more than 100 participants, we include the study.

**No: not a study which reports mental health symptom changes longitudinally pre-COVID-19 to COVID-19 or during COVID-19.** If the study does not report continuous scores of symptom levels or proportions of participants meeting the threshold on a validated scale, or diagnostic criteria using a validated diagnostic interview prior to and after the start of COVID-19, or longitudinally during COVID-19, then it will be excluded.

For pre-COVID versus during-COVID studies, pre- and during- samples must include the same cohort, not different representative samples. Pre- and during-samples should have less than 10% difference in the participants in the sample* or should statistically account for missing data, i.e., if N between the samples differs by more than 10%, modelling or imputation is needed to evaluate results for all participants. Pre-COVID data needs to be collected prior to 2020 (or at least 80% of the participants’ data need be collected prior to 2020 if collection spans from 2019 to 2020) and after 2018 (or at least 80% of the participants’ data need to be collected after 2018 if collection spans from pre-2018 to 2018).

For studies with multiple waves across COVID, if there are pre-pandemic time points, the most recent pre-pandemic wave needs to be in 2018 or later; if the most recent pre-pandemic wave spans from pre-2018 to 2018, at least 80% of the data need to be collected in 2018. Studies with multiple waves across COVID-19 must have at least two time points that have less than 10% difference in the participants in the sample*, or should statistically account for missing data, regardless of whether or not the study has pre-COVID assessments.

If outcomes from the study are only shown graphically without eligible numerical values, exclude the study.

* At least 90% of participants in assessments from two time points need to be the same participants. In a three-wave survey, if N-T1 = 1000, N – T2 = 500, and N – T3 = 500, T2 and T3 would only be eligible if at least 90% of the participants at each time point were the same. It is not enough to just have a total N within 10%.

**Yes: study eligible for inclusion in systematic review.**

**Appendix 3: Adequacy of Study Methods and Reporting**

**Q1. Was the sample frame appropriate to address the target population?**

**Yes:** The sampling frame was a true or close representation of the target population.

**No:** The sampling frame was NOT a true or close representation of the target population.

**Unclear:** Not enough information provided to determine.

**Q2. Were study participants recruited in an appropriate way?**

**Yes:** A census was undertaken, OR, some form of random selection was used to select the sample (e.g. simple random sampling, stratified random sampling, cluster sampling, systematic sampling).

**No:** A census was NOT undertaken, AND some form of random selection was NOT used to select the sample.

**Unclear:** Not enough information provided to determine.

**Q3. Was the sample size adequate?**

**Yes:** There is evidence that the authors conducted a sample size calculation to determine an adequate sample size OR the study was large enough (e.g., a large national survey) whereby a sample size calculation is not required. In these cases, sample size can be considered adequate. If at least 200 participants are included for continuous outcomes and 250 for proportions, this is considered low risk.

**No:** The authors did not reach their intended sample size, or no sample size calculation is provided and there are < 100 participants for continuous outcomes, or < 125 for proportions.

**Unclear:** No sample size calculation is provided, and between 100-199 participants are included for continuous outcomes or between 125-249 for proportions.

**Q4. Were the study participants and setting described in detail?**

**Yes:** Data included age, sex, and at least 1 socioeconomic indicator (e.g., income, education, work status).

**No:** The minimum sociodemographic variables have not been reported.

**Unclear:** Not stated

**Q5. Was the response rate adequate and was the data analysis conducted with sufficient coverage?**

**Yes:** The overall response rate or response rate for intended subgroups was >/=75%, OR, an analysis was performed that established that there was not a substantive difference in relevant demographic characteristics between responders and non-responders within a subgroup (if non-response too high (e.g., > 50%), code “No”)

**No:** The overall response rate or response rate for subgroups was <75%, and if any analysis comparing responders and non-responders was done, it showed a meaningful difference in relevant demographic characteristics between responders and non-responders.

**Unclear:** Not enough information provided to determine.

**Q6. Were valid methods used for the identification of the outcome variable?**

**Yes:** The study instrument had been shown to have reliability and validity, e.g., test-retest, piloting, validation in a previous study, etc.

**No:** The study instrument had NOT been shown to have reliability or validity.

**Unclear:** Not stated.

**Q7. Was the mental health outcome measured in a standard, reliable way for all participants?**

**Yes:** All self-report data were collected directly from the participants. Any clinical interview data includes at least information about the interviewers’ level of education or training received. The same mode of data collection was used for all participants. All aspects of this question must be present (where relevant).

**No:** In some instances, data were collected from a proxy (e.g., a spouse). The qualifications of clinical interviewers are not reported or not appropriate. The same mode of data collection was NOT used for all participants. If any aspects of this item are absent, it is high risk.

**Unclear**: Not stated.

**Q8. Was there appropriate statistical analysis?**

**Yes:** Continuous variables report (1) mean (SD) of change or (2) pre mean (SD) and post mean (SD) with/out correlation between pre and post scores. For dichotomous variables, numerator, denominator, and percentages are clearly reported. Continuous variables are not artificially dichotomized. The statistical analyses section is detailed enough for readers to understand change scores (see STROBE reporting guidelines, if necessary).

**No:** Continuous variables do not include a report of the (1) mean (SD) of change or (2) pre mean (SD) and post mean (SD) with/out correlation between pre and post scores. For dichotomous variables, the numerator, denominator, or percentages are not clearly reported. The statistical analyses section does not clearly describe the methods used to assess change scores.

**Q9. Was the follow-up rate adequate, and if not, was the low follow-up rate managed appropriately?**

**Yes:** At least 75% of those who participated in the pre-COVID-19 assessment(s) provided follow-up responses and had their responses included in the follow-up, OR, an analysis was performed that showed no substantive difference in relevant demographic characteristics between participants who stayed in the study and drop-outs (if dropout too high (e.g. > 50%), code “No”).

**No:** Less than 75% of those participated in the pre-COVID-19 assessment(s) provided responses and had their responses included in the follow-up, and if any analysis comparing participants who stayed in the study and drop-outs was done, it showed a substantive difference in relevant demographic characteristics between the two groups.

**Unclear:** Not stated.

**Appendix 4. Confidence Intervals for Proportion Change When Using 30% and 70% of Pre-COVID-19 Cases were Cases During COVID-19: Calculated for Studies that Did Not Provide 95% Confidence Intervals for Change**

| **First Author** | **Outcome Domain** | **Sex/Gender** | **95% CI with 30%** | **95% CI with 70%** |
| --- | --- | --- | --- | --- |
| Dong^55^ | General Mental Health | Females/Women | -0.10, -0.06 | -0.10, -0.07 |
|  |  | Males/Men | -0.10, -0.04 | -0.10, -0.04 |
| Saraswathi^56^ | Anxiety Symptoms | Females/Women | -0.24, -0.03 | -0.22, -0.06 |
|  |  | Males/Men | -0.23, 0.06 | -0.20, 0.02 |
|  | Depression Symptoms | Females/Women | -0.10, 0.14 | -0.05, 0.09 |
|  |  | Males/Men | -0.25, 0.05 | -0.21, 0.01 |
|  | Stress | Females/Women | -0.12, 0.06 | -0.09, 0.04 |
|  |  | Males/Men | -0.20, 0.08 | -0.16, 0.03 |
| van der Velden^49^ | General Mental Health | Females/Women | -0.02, 0.03 | -0.01, 0.02 |
|  |  | Males/Men | -0.03, 0.01 | -0.02, 0.00 |
| van der Velden^50^ | General Mental Health | Females/Women | -0.01, 0.03 | -0.00, 0.03 |
|  |  | Males/Men | -0.03, 0.01 | -0.03, 0.00 |

**Appendix 5. PRISMA Flowchart**

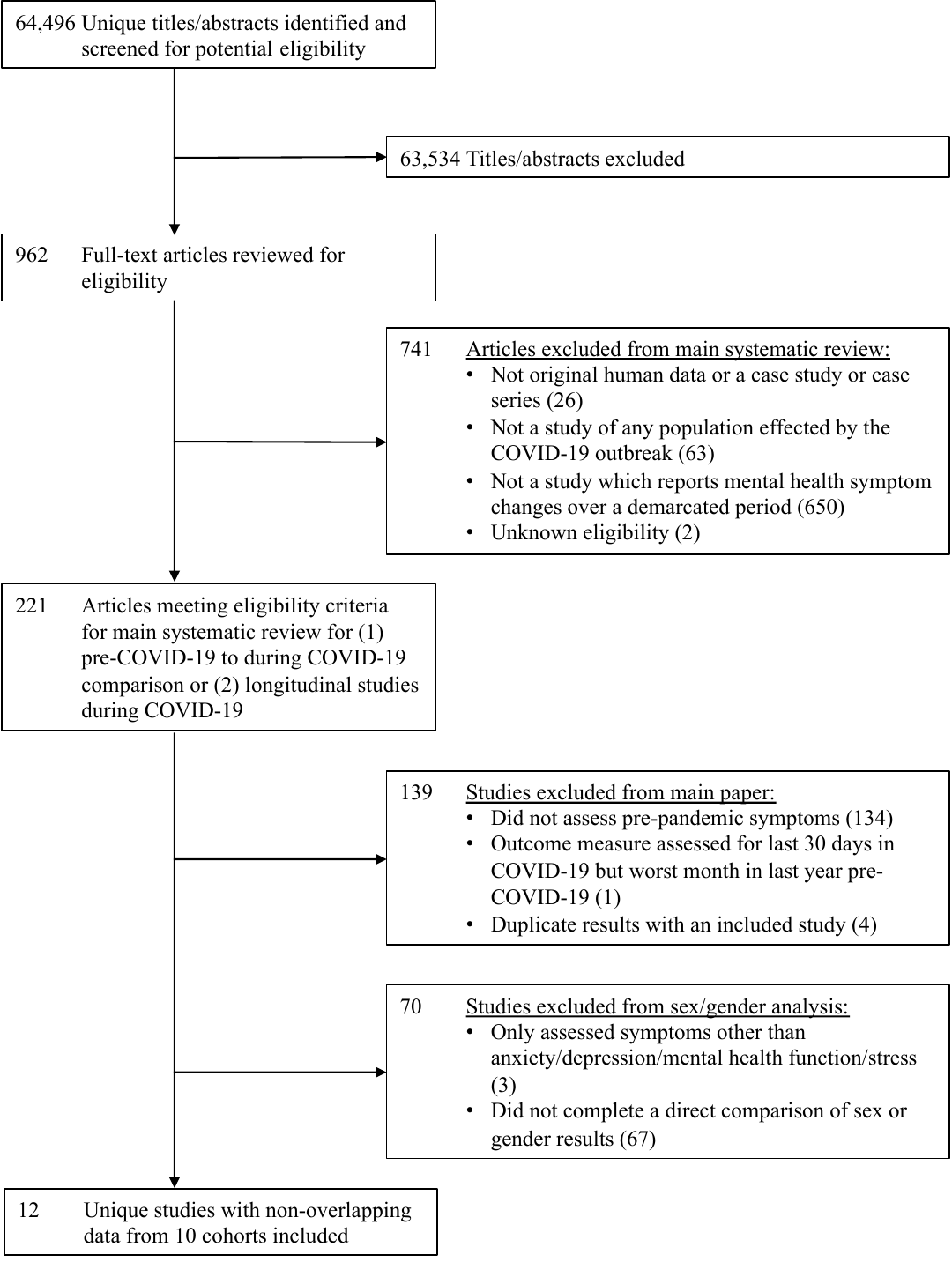

**Appendix 6. Adequacy of Methods and Reporting of Included Studies (N=12)**

| **Author** | **Appropriate**  **sample**  **frame** | **Appropriate participant recruitment** | **Adequate**  **sample size** | **Subjects**  **and setting adequately described** | **Adequate response rate and data analysis with sufficient coverage** | **Valid methods for identification of outcome variable** | **Standard, reliable outcome measurement** | **Appropriate statistical analysis** | **Adequate follow-up response rate/ appropriate management of low response rate** |
| --- | --- | --- | --- | --- | --- | --- | --- | --- | --- |
| Dong^55^ | No | No | Yes | Yes | Yes | Yes | Yes | Yes | Yes |
| Lim^58^ | Yes | Unclear | Yes | Yes | Unclear | Yes | Unclear | Yes | Unclear |
| Magson^54^ | No | Unclear | Yes | Yes | Unclear | Yes | Yes | Yes | No |
| Megías-Robles^51^ | Unclear | No | Unclear | No | Unclear | Yes | Yes | Yes | No |
| Pierce^47^ | Yes | Yes | Yes | Yes | Unclear | Yes | Yes | Yes | Unclear |
| Daly^48^ |  |  |  |  |  |  |  |  | No |
| Rimfeld^52^ | Yes | Unclear | Yes | Yes | Unclear | Yes | Yes | Yes | Unclear |
| Saraswathi^56^ | No | Yes | Unclear | Yes | Yes | Yes | Yes | Yes | Yes |
| Savage^57^ | No | Yes | Yes | Yes | No | Yes | Yes | Yes | No |
| Shanahan^53^ | No | Unclear | Yes | Yes | Unclear | Yes | Yes | Yes | Yes |
| van der Velden^49^ | Yes | Yes | Yes | Yes | Yes | Yes | Yes | Yes | Yes |
| van der Velden^50^ |  |  |  |  |  |  |  |  |  |
